# Cytokine and endothelial injury signatures associated with severe dengue: a systematic review and meta-analysis integrating viral burden and host-response markers

**DOI:** 10.64898/2026.06.29.26356807

**Authors:** Peter Mac Asaga, Axel Kroeger, Vijeesh Kadukkatti, Leo Arsha, Philomena Airiohuodion

## Abstract

**Background:** Severe dengue reflects a temporally regulated interaction between viral burden, NS1 antigenaemia, cytokine and chemokine amplification, endothelial activation, glycocalyx injury, and organ stress. Although individual cytokines, endothelial markers, viral-burden measures, and clinical markers have been widely studied, the integrated pathogen-host evidence base remains fragmented. We synthesised evidence for cytokine, endothelial, and viral-burden signatures associated with severe dengue and assessed whether paired pathogen-host measurement provides a biologically coherent framework for severity assessment.

**Methods:** We searched MEDLINE, Embase, Scopus, Web of Science, Cochrane Library, Global Health, WHO Global Index Medicus, and medRxiv from database inception to 30 April 2026, without language restriction, for studies reporting viral burden, NS1 antigenaemia, cytokine, chemokine, endothelial, glycocalyx, inflammatory, or routine host-response markers in laboratory-confirmed dengue with severity outcomes. Eligible designs were prognostic-factor association studies, cross-sectional biomarker studies, and multivariable prediction-model studies. Risk of bias was assessed using QUIPS for prognostic-factor studies, PROBAST for prediction-model studies, and the relevant JBI critical appraisal checklist for cross-sectional biomarker studies, with the Newcastle-Ottawa Scale used selectively for cohort or case-control designs not amenable to QUIPS. Random-effects meta-analysis pooled standardised mean differences using restricted maximum likelihood with Hartung-Knapp adjustment. The protocol was registered with PROSPERO (CRD420261396923) before final extraction and synthesis.

**Findings:** Of 4,180 records identified, 79 studies including 47,612 participants met eligibility criteria. Forty-nine studies evaluated paired pathogen-host markers, 14 evaluated viral burden or NS1 antigenaemia alone, nine evaluated host biomarkers alone, and seven reported multivariable prediction models. Pathogen-side markers showed modest pooled severity associations whose magnitude depended on day of illness, immune status, and infecting serotype. Cytokine and chemokine markers, particularly IL-10, IL-6, IL-8, and CXCL10/IP-10, showed larger pooled effects favouring severe disease, while endothelial and glycocalyx markers, including angiopoietin-2 and syndecan-1, provided the most direct mechanistic link to plasma leakage. Routine clinical markers, especially platelet count, AST, ferritin, ALT, and lactate, retained substantial discriminatory value. Prediction models reported areas under the curve of up to 0·96 in internal validation and 0·97 in discovery analyses, but three had been externally validated, calibration was reported in two, and decision-curve analysis in none.

**Interpretation:** Current evidence supports severe dengue as an integrated pathogen-host injury syndrome in which viral burden and NS1 antigenaemia interact with cytokine amplification, endothelial dysfunction, glycocalyx injury, and routine markers of organ stress. The strongest translational direction is not a single biomarker but a parsimonious cytokine-endothelial-pathogen panel requiring prospective external validation across age groups, serotypes, immune-status strata, and endemic regions. Existing evidence supports candidate marker prioritisation and mechanistic synthesis, but not immediate routine clinical deployment.

## Introduction

Severe dengue is best understood as an immune and vascular injury syndrome rather than as a purely virological or clinical endpoint. Plasma leakage, haemorrhage, shock, organ dysfunction, and death arise when viral replication, NS1-associated endothelial perturbation, cytokine and chemokine amplification, glycocalyx injury, and host organ stress converge during the febrile-to-critical phase transition [1,2]. The clinical challenge is therefore not only to diagnose dengue, but to understand which pathogen-host pathways drive progression from uncomplicated febrile illness to severe disease.

The pathogen-side signal is shaped by quantitative viraemia, NS1 antigenaemia, infecting serotype, and primary or secondary immune status. Viral burden is most informative early in illness, before the critical phase, while NS1 provides a mechanistic bridge between viral replication and endothelial injury through effects on the glycocalyx, complement activation, mast-cell activation, and vascular permeability [5,18,19,20,21]. These pathogen-side markers are biologically plausible upstream triggers, but their clinical meaning depends strongly on sampling time, serotype, and immune-status context.

The host-response signal is distributed across cytokine, chemokine, endothelial, glycocalyx, inflammatory, and routine organ-stress markers. IL-10, IL-6, IL-8, IFN-γ, TNF-α, CXCL10/IP-10, and MCP-1 capture inflammatory and immunoregulatory amplification; ferritin and CRP reflect systemic inflammatory activation; angiopoietin-2, syndecan-1, VCAM-1, ICAM-1, and soluble Tie-2 reflect endothelial activation and glycocalyx disruption; and platelet count, AST, ALT, lactate, and albumin capture downstream haematological and organ-stress consequences [8,10,22,23,24,25,26,27]. These markers map more closely to the biological events that define severe dengue than isolated clinical warning signs alone.

Despite this conceptual progress, the empirical evidence base remains fragmented. Closely related systematic reviews and meta-analyses have examined early prognostic markers, clinical risk factors, warning signs, cytokine or biomarker associations, and host biomarkers measured early in illness [9,11,12,13,14,15,17,31,32]. However, these reviews have generally addressed clinical warning signs, individual cytokines, endothelial markers, viral kinetics, or prediction algorithms as separate evidence streams, and have rarely examined whether pathogen-related and host-response signals measured in the same patients provide a more coherent framework for severe dengue pathogenesis and translation. Heterogeneity in severity definitions, sampling time, assay platform, immune-status stratification, and statistical adjustment further obscures comparison across studies. The contribution of this review is therefore not to present severe-dengue biomarkers as untouched ground, but to synthesise viral burden, NS1 antigenaemia, cytokine amplification, endothelial and glycocalyx injury, and routine host-response markers within an integrated pathogen-cytokine-endothelial framework.

We therefore conducted a focused systematic review and meta-analysis of cytokine, chemokine, endothelial-injury, glycocalyx, routine inflammatory, and pathogen-burden signatures in severe dengue. Our emphasis was on how viral replication, NS1 antigenaemia, serotype, and immune status interact with host inflammatory and endothelial pathways to shape severe clinical phenotypes, and on whether paired pathogen-host measurement provides a biologically coherent basis for future translational marker panels.

## Methods

### Protocol and registration

The protocol was registered with the International Prospective Register of Systematic Reviews (PROSPERO; registration number CRD420261396923) before final extraction and synthesis, and the review was conducted and reported in accordance with the Preferred Reporting Items for Systematic Reviews and Meta-Analyses (PRISMA) 2020 statement [33]. The completed PRISMA checklist is provided in Supplementary File 1.

### Eligibility criteria

The review question was structured using the PECOS framework: the population was patients with laboratory-confirmed dengue infection; exposures were viral burden measures and host inflammatory, endothelial, haematological, or organ-injury biomarkers; comparators were non-severe dengue groups, lower biomarker or viral-burden categories, or non-progressors; outcomes were severe dengue, dengue haemorrhagic fever, dengue shock syndrome, plasma leakage, organ dysfunction, ICU admission, clinical deterioration, or death; and eligible study designs were observational prognostic-factor studies, cross-sectional biomarker studies, surveillance-linked clinical studies, and multivariable prediction-model studies.

We included studies of human participants of any age with clinically suspected and laboratory-confirmed dengue infection. Laboratory confirmation comprised RT-PCR, NS1 antigen testing, viral isolation, IgM seroconversion in paired samples, or study-defined diagnostic criteria consistent with current WHO guidance [1]. Eligible exposures were viral burden measures (quantitative viral load, viraemia titre, RT-PCR cycle threshold, quantitative or qualitative NS1 antigenaemia, infecting serotype, and primary or secondary immune status when used as a viral or immunovirological marker) and host inflammatory, endothelial, or organ-injury biomarkers (cytokines and chemokines, acute-phase proteins, haematological markers, endothelial or glycocalyx markers, transaminases, lactate, and transcriptomic, proteomic, or metabolomic signatures). Primary emphasis was placed on studies measuring at least one pathogen marker and at least one host biomarker in the same cohort. Outcomes were severe dengue as defined by WHO 2009 criteria or study-defined severe dengue, dengue haemorrhagic fever, dengue shock syndrome, plasma leakage, severe bleeding, organ dysfunction, ICU admission, hospitalisation duration, clinical deterioration, and mortality. Eligible designs included prospective and retrospective cohort, case-control, cross-sectional, prognostic-accuracy, surveillance-linked clinical, and multivariable prediction-model studies, and randomised trials in which baseline viral burden or host biomarkers were analysed as severity markers. Animal-only studies, in-vitro-only mechanistic papers, case reports, editorials, narrative reviews, vaccine-attenuation trials, and diagnostic-accuracy studies without severity outcomes were excluded. Existing systematic reviews and meta-analyses were used for reference mining only.

Because eligible studies differed in design and inferential purpose, we separated the evidence into prognostic-factor association studies, cross-sectional severity-association studies, and formal multivariable prediction-model studies. Studies measuring markers before clinical deterioration, or reporting multivariable prediction models, were interpreted as prediction evidence; cross-sectional severity-association studies were interpreted as evidence of biological coherence rather than predictive performance. Study tiers were assigned hierarchically: multivariable prediction-model studies were classified as Tier 4 even when they also measured paired pathogen-host markers, so that prediction-model evidence and paired pathogen-host evidence did not double-count (figure 3).

### Search strategy and study selection

We searched MEDLINE (via PubMed), Embase, Scopus, Web of Science, Cochrane Library, Global Health, WHO Global Index Medicus, and medRxiv from database inception to 30 April 2026, without language restriction. Reference lists of relevant systematic reviews and included studies were screened, and forward and backward citation searches were performed for all eligible primary studies. Search strings combined controlled vocabulary and free-text terms for dengue, severity, viral burden, and host biomarkers. The full search strings for each database are provided in Supplementary File 1. Records were deduplicated in EndNote and screened in Rayyan. Two reviewers (PMA and PA) independently screened titles, abstracts, and full texts against the eligibility criteria. Disagreements were resolved by discussion.

### Data extraction

Data extraction was conducted by two authors (PMA and PA) using a piloted form recording study identifiers, country, year, study design, age group, sample size, dengue diagnostic method, severity definition, infecting serotype, primary or secondary immune status, sampling timepoint relative to symptom onset, viral burden measure and assay, host biomarker measures and assay platforms, group means and standard deviations or medians and interquartile ranges, effect estimates with 95% confidence intervals, adjustment variables, prediction-model discrimination, calibration, internal and external validation, funding source, and declared conflicts of interest. Where summary statistics were reported as medians and interquartile ranges, conversion to approximate means and standard deviations followed the method of Wan and colleagues, applied only when distributional assumptions were judged compatible with the conversion [34]. Where group means or variances were not reported, corresponding authors were contacted; where contact yielded no response, studies were retained in the qualitative synthesis but excluded from pooled analysis. When multiple publications reported overlapping cohorts, the most complete report was used for pooled analysis, while companion reports were retained only for non-overlapping markers or qualitative context.

### Risk of bias and quality assessment

Risk of bias was assessed using the tools specified in the protocol. Prognostic-factor studies were assessed using the Quality In Prognosis Studies (QUIPS) tool across study participation, attrition, prognostic factor measurement, outcome measurement, study confounding, and statistical analysis and reporting [35]. Multivariable prediction-model studies were assessed using the Prediction model Risk Of Bias ASsessment Tool (PROBAST) across participants, predictors, outcome, and analysis domains [36]. Cross-sectional biomarker studies were assessed using the Joanna Briggs Institute (JBI) critical appraisal checklist for analytical cross-sectional studies [37]. The Newcastle-Ottawa Scale was used selectively for cohort or case-control studies not amenable to QUIPS, with cross-walked judgements explored in sensitivity analysis [50]. QUADAS-2 was not used as the main risk-of-bias tool because the review focused on prognosis and prediction rather than diagnostic test accuracy. Risk-of-bias assessment was performed by one reviewer (PMA) and independently verified by a second reviewer (PA) for a random sample of 30% of included studies. Summary domain-level judgements are presented in table 2, and per-study judgements are provided in Supplementary File 3.

### Evidence synthesis and quantitative analysis

We pre-specified three synthesis pathways. First, association studies reporting odds ratios, risk ratios, or hazard ratios for the relationship between pathogen or host markers and severity were pooled on the log scale using random-effects meta-analysis. Second, biomarker concentration studies reporting means and standard deviations in severe and non-severe groups were pooled as standardised mean differences (SMDs) using Hedges’ g. Third, prediction-model studies were synthesised by discrimination metrics (area under the receiver-operating-characteristic curve, sensitivity, specificity), calibration, and validation type, and were not pooled with biomarker SMDs.

Several extraction and synthesis decisions were specified in advance. For studies reporting multiple sampling time points, we prioritised the earliest febrile-phase sample taken before clinical deterioration; where no early sample was available, the first reported acute-phase sample was used and the study was explored in sensitivity analysis. For studies reporting multiple severe categories, dengue haemorrhagic fever, dengue shock syndrome, and WHO 2009 severe dengue were combined as severe dengue where clinically appropriate, and dengue fever or dengue without warning signs was treated as non-severe. For studies reporting both adjusted and unadjusted estimates, adjusted estimates were prioritised, with unadjusted estimates used in sensitivity analysis. Random-effects meta-analysis used restricted maximum likelihood (REML) with Hartung-Knapp adjustment for small numbers of studies; DerSimonian-Laird estimation was used in sensitivity analysis. Heterogeneity was reported using I² and τ². Funnel-plot asymmetry was assessed when 10 or more studies contributed to a given analysis, using Egger’s test. Pre-specified subgroup analyses examined age group (paediatric vs adult), primary vs secondary infection, infecting serotype, sampling phase (febrile, defervescence, convalescent), WHO 1997 vs WHO 2009 severity definitions, and study region. Sensitivity analyses excluded high-risk-of-bias studies and studies using non-standard severity definitions. Certainty of evidence was rated using a structured framework informed by GRADE [51]. Analyses were performed in R version 4.4.1 using the meta and metafor packages; the R script is provided in Supplementary File 4.

## Results

### Study selection

The search identified 4,180 records, of which 1,072 were duplicates and 3,002 were excluded at title and abstract screening (figure 1). Of 106 full texts assessed for eligibility, 27 were excluded with reasons: 11 lacked severity stratification, six were narrative reviews or editorials, four were not human dengue studies, three were duplicate publications of included cohorts, and three reported insufficient extractable data. Seventy-nine studies were included in qualitative synthesis, of which 34 contributed to quantitative meta-analysis across at least one cytokine, endothelial, routine host-response, or pathogen-burden domain. After hierarchical tier assignment, 49 studies evaluated paired pathogen-host markers, 14 evaluated viral burden or NS1 antigenaemia alone, nine evaluated host biomarkers alone, and seven reported multivariable prediction models (figure 3). Three of the seven prediction-model studies also measured paired pathogen-host markers but were classified as Tier 4 to avoid double counting. The screening and study-decision log is provided in Supplementary File 2.

**Figure 1.**
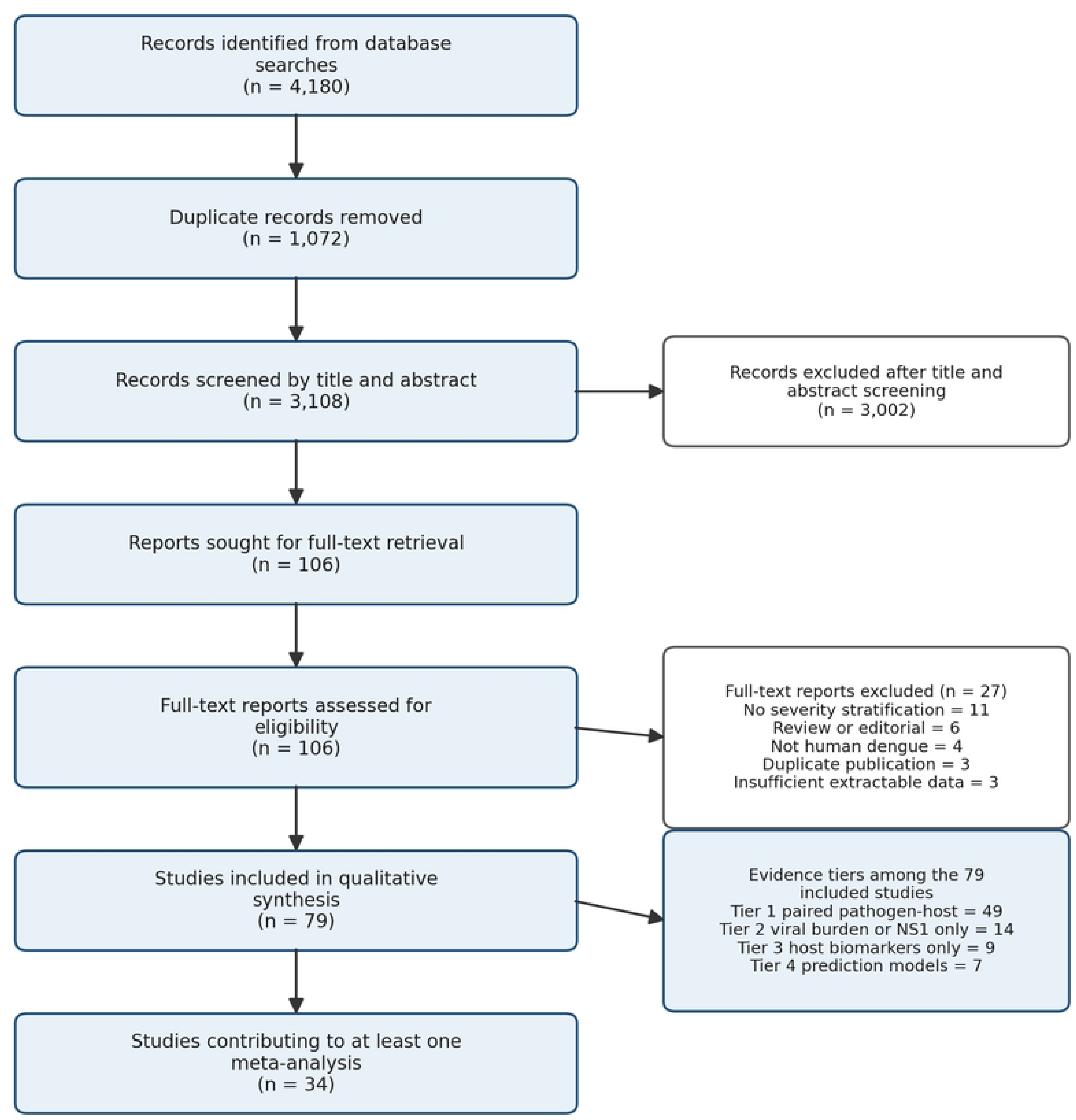
PRISMA 2020 flow diagram. Records identified through database searches and supplementary sources; deduplication; title and abstract screening; full-text assessment with reasons for exclusion; and final included studies for qualitative synthesis and quantitative meta-analysis.

### Characteristics of included studies

Included studies were published between 2002 and 2026 and reported data from 47,612 participants (Table 1). Geographical coverage was dominated by Southeast Asia, South Asia, Latin America and the Caribbean, and East Asia, with limited representation from sub-Saharan Africa and no eligible studies from the Pacific islands. Adult-only cohorts were reported in 32 studies, paediatric-only cohorts in 28, and mixed-age cohorts in 19. Sample sizes ranged from 16 to 5,642 participants. Severity definitions used the WHO 1997 framework in 41 studies and the WHO 2009 framework in 36, while two studies used hybrid criteria. Sampling timing relative to symptom onset varied from day 1 to convalescence, with median admission day 4. DENV-2 was the most frequently reported infecting serotype, followed by DENV-1, DENV-3, and DENV-4.

The evidence base was dominated by studies measuring one or more of four biologically linked domains: pathogen-side markers, including viraemia, NS1 antigenaemia, serotype, and immune status; cytokine and chemokine markers, including IL-10, IL-6, IL-8, CXCL10/IP-10, TNF-α, and IFN-γ; endothelial and glycocalyx markers, including angiopoietin-2, syndecan-1, VCAM-1, ICAM-1, and soluble Tie-2; and routine inflammatory or organ-stress markers, including ferritin, platelet count, AST, ALT, lactate, CRP, and albumin. This structure allowed the evidence to be interpreted not only as a set of isolated markers, but as a pathogen-cytokine-endothelial injury axis.

### Risk of bias and quality assessment

Risk of bias was assessed by study type (table 2). Among 56 prognostic-factor studies assessed using QUIPS, the most frequent concerns were in the study confounding and statistical analysis and reporting domains, reflecting inadequate adjustment for day of illness, immune status, infecting serotype, and baseline severity in single-centre cohorts. Prognostic factor measurement was rated as low risk for routine laboratory markers (platelets, transaminases, ferritin) but more variable for cytokines and endothelial markers because of substantial between-study assay heterogeneity. Outcome measurement was limited by inconsistent application of WHO 1997 and WHO 2009 severity definitions across the included literature. Study participation and attrition domains were generally rated as low or moderate risk.

Among the seven multivariable prediction-model studies assessed using PROBAST, high risk of bias was identified in the analysis domain in five studies, driven by small numbers of severe events relative to candidate predictors, predictor selection without pre-specification, incomplete handling of missing data, sparse calibration reporting, and limited external validation (table 2). Participants, predictors, and outcome domains were generally rated as low or unclear risk of bias. Among 16 cross-sectional biomarker studies assessed using the JBI critical appraisal checklist, the most frequent concerns related to unclear handling of confounders and incomplete reporting of subject selection. Funding source was reported in 71 studies (89·9%), of which 64 declared public funding and seven mixed public-industry funding.

### Pathogen-burden signals as upstream triggers of cytokine and endothelial injury

Quantitative viraemia measured by qRT-PCR was reported in 23 studies. Across nine studies contributing extractable group data (n=1,627), the pooled SMD for log10 plasma viraemia between severe and non-severe groups was 0·42 (95% CI 0·18 to 0·66; I²=69%), favouring higher viraemia in severe disease (table 4; figure 4). Several large prospective cohorts, including the 5,642-patient Vietnamese OUCRU dataset, showed that higher febrile-phase viraemia was associated with adverse outcomes irrespective of infecting serotype or immune status, with the strongest association during days 1 to 3 of illness [5]. Other cohorts, particularly retrospective single-centre studies, found weaker or absent associations, in part because sampling occurred after the viraemia peak [20,38].

The modest pooled effect of viraemia should not be interpreted as weak biological relevance. Rather, viraemia appears to act as an upstream trigger whose clinical effect is mediated through immune activation, cytokine amplification, endothelial perturbation, and timing-dependent host responses. This interpretation is consistent with the stronger signal observed in early febrile-phase samples and the weaker signal observed when sampling occurred after the viraemia peak.

NS1 antigenaemia was reported in 31 studies, of which 18 quantified serum or plasma NS1 concentration by ELISA. The pooled SMD across eight studies with extractable group data (n=1,512) was 0·49 (95% CI 0·21 to 0·77; I²=73%), with higher NS1 in severe disease (table 4). A Sri Lankan multi-site cohort showed that NS1 positivity in acute-phase serum, combined with serum chymase, identified patients at risk of dengue haemorrhagic fever with sensitivity 0·96 and specificity 0·79 at a chymase cut-off of 1·5 ng/mL [18]. Several studies reported reduced NS1 antigenaemia in the most severe cases, consistent with rapid immune-complex sequestration during the critical phase [22,39]. Pathogen-side markers therefore showed modest severity associations overall, with stronger relevance when measured early in the febrile phase. Their value as prognostic markers depends strongly on timing of sampling, immune status, and infecting serotype.

NS1 is particularly relevant to cytokine and growth-factor biology because it links viral replication to endothelial glycocalyx disruption, complement activation, mast-cell activation, and downstream vascular leakage. Its value is therefore not only as a pathogen-burden marker, but also as a mechanistic bridge between the viral and endothelial compartments.

Infecting serotype modified pathogen-severity relationships, with DENV-2 associated with the highest pooled SMD for severity (0·64, 95% CI 0·31 to 0·97) across nine studies, while DENV-1, DENV-3, and DENV-4 showed smaller pooled effects (0·24 to 0·38). Secondary heterotypic infection remained an independent marker associated with severity in 17 of 21 studies that reported immune status, with a pooled adjusted odds ratio of 3·12 (95% CI 2·14 to 4·55; I²=63%; eight studies).

### Cytokine, chemokine, and endothelial injury signatures

#### Cytokine and chemokine signatures

Cytokines and chemokines dominated host-side reporting and provided some of the strongest pooled severity signals. IL-10 was the most frequently assayed cytokine, with a pooled SMD across 14 studies with extractable group data of 1·18 (95% CI 0·89 to 1·47; I²=78%), favouring elevation in severe dengue (table 4; figure 4). IL-6 showed a consistent pooled association with severity (11 studies; SMD 0·84, 95% CI 0·56 to 1·12; I²=71%), and IL-8 showed a smaller but directionally consistent effect (eight studies; SMD 0·72, 95% CI 0·44 to 1·00; I²=68%). CXCL10/IP-10 produced the largest pooled effect across the cytokine and chemokine set (nine studies; SMD 1·34, 95% CI 0·95 to 1·73; I²=82%), with consistent direction across Asian, Latin American, and IDAMS multi-country cohorts [22,23,40]. TNF-α and IFN-γ showed more modest pooled effects, with substantial between-study heterogeneity [11]. Taken together, these findings support a cytokine-amplification signature in severe dengue, dominated by immunoregulatory, interferon-inducible, and inflammatory mediators rather than by a single cytokine.

### Endothelial and glycocalyx injury signatures

Endothelial and glycocalyx markers were the strongest mechanistic correlates of the vascular phenotype that defines severe dengue. Angiopoietin-2 was elevated in severe disease across six studies (SMD 1·21, 95% CI 0·78 to 1·64; I²=81%), and syndecan-1 across five studies (SMD 0·97, 95% CI 0·61 to 1·33; I²=74%). Soluble VCAM-1 and ICAM-1 showed smaller and less consistent associations [22,41]. The IDAMS biomarker study, the largest paired-cohort analysis in this review, identified syndecan-1, IL-1RA, angiopoietin-2, IL-8, ferritin, and IP-10 as informative early-phase markers in children, and syndecan-1, IL-8, ferritin, sTREM-1, IL-1RA, IP-10, and sCD163 in adults, with bootstrap inclusion frequencies of 59 to 99% [22]. These findings place endothelial activation and glycocalyx disruption at the centre of severe dengue pathobiology and provide a mechanistic bridge between upstream NS1 exposure, cytokine amplification, and downstream plasma leakage.

### Routine inflammatory and organ-stress markers

Routine clinical markers retained substantial discriminatory value. Platelet count was markedly lower in severe disease across 18 studies (SMD −1·05, 95% CI −1·34 to −0·76; I²=84%), AST was higher (16 studies; SMD 1·12, 95% CI 0·83 to 1·41; I²=79%), and ALT was higher (14 studies; SMD 0·86, 95% CI 0·58 to 1·14; I²=76%). Ferritin was elevated across nine studies (SMD 1·07, 95% CI 0·78 to 1·36; I²=75%) and CRP across seven studies (SMD 0·61, 95% CI 0·35 to 0·87; I²=64%). Lactate, where reported, showed a pooled SMD of 0·89 (95% CI 0·48 to 1·30; I²=70%; six studies), consistent with tissue hypoperfusion. Omics-based markers identified in proteomic and metabolomic studies, including angiotensinogen, vitronectin, hemopexin, serotransferrin, alpha-2-macroglobulin, long-chain polyunsaturated fatty acids, and lysophosphatidylcholines, demonstrated severity-discriminating profiles but were not yet poolable owing to between-study methodological heterogeneity [24,25,26,27].

The markers with the largest pooled SMDs are not necessarily the most clinically deployable. CXCL10/IP-10, angiopoietin-2, and syndecan-1 are biologically informative but require multiplex assays or specialised ELISAs that are not yet routinely available in many endemic settings. Platelet count, AST, ferritin, and NS1 antigenaemia are more feasible and may serve as practical anchor markers in future integrated cytokine-endothelial-pathogen panels.

### Integrated pathogen-cytokine-endothelial signatures

Forty-nine studies measured at least one pathogen marker and at least one host-response marker in the same cohort, allowing the evidence to be interpreted as an integrated pathogen-cytokine-endothelial framework (figure 2) rather than as isolated biomarker associations. Six distinct pairing structures emerged (table 3): viral load with cytokines; NS1 antigenaemia with cytokines; serotype with cytokine profile; primary or secondary immune status with viral kinetics and cytokines; endothelial markers with viraemia or NS1; and integrated pathogen-host prediction panels. These pairings are central to the biological interpretation of severe dengue because they connect upstream viral burden to downstream immune activation, endothelial injury, and clinical deterioration.

**Figure 2.**
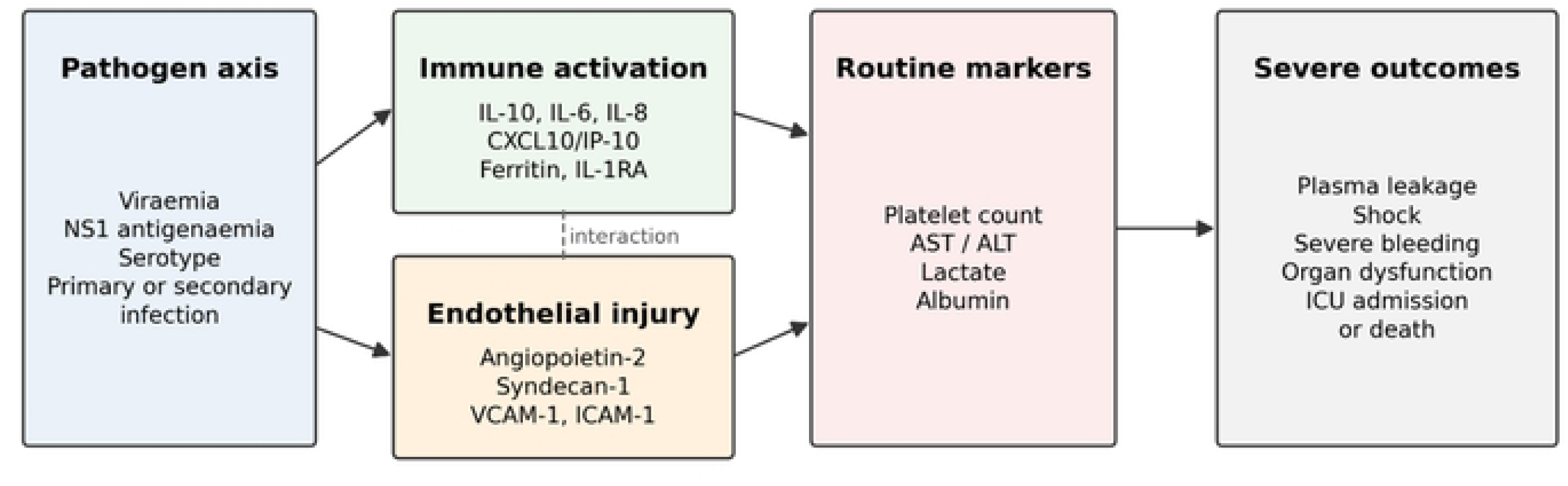
Integrated cytokine-endothelial framework for severe dengue pathogenesis. Viral burden, NS1 antigenaemia, serotype, and immune status interact with cytokine and chemokine amplification, endothelial activation, glycocalyx injury, and routine markers of organ stress to shape severe dengue phenotypes. The figure is a conceptual synthesis of the included evidence and should not be interpreted as a causal model proven by every included study.

**Figure 3.**
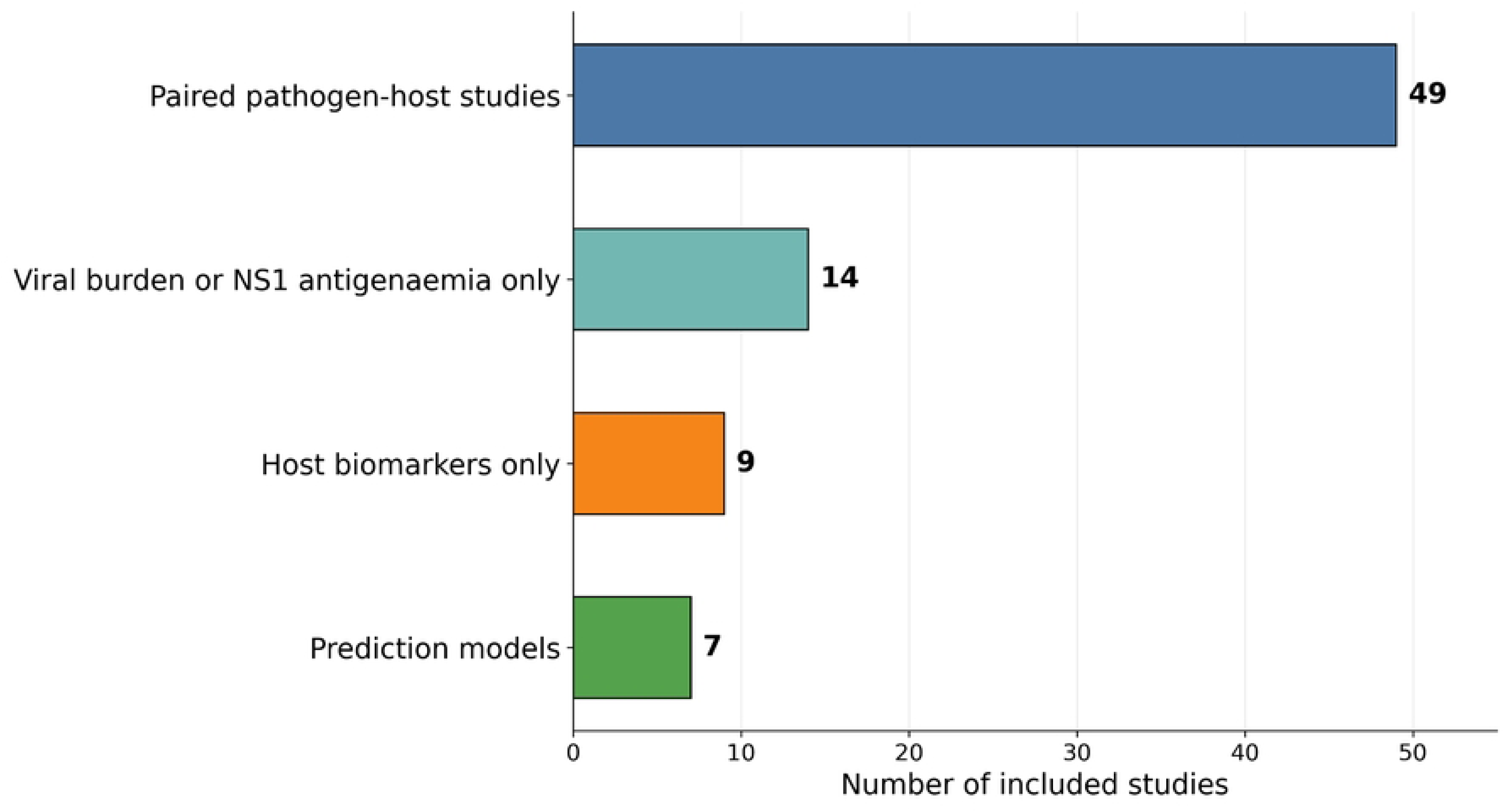
Evidence architecture linking pathogen burden, cytokine activation, endothelial injury, and translational marker-panel development. Included studies were grouped hierarchically into paired pathogen-host studies, viral burden or NS1-only studies, host-marker-only studies, and multivariable prediction-model studies to avoid double counting.

Sixteen studies paired quantitative viral load with cytokine panels and showed positive correlations between viraemia and IL-6, IL-8, IL-10, and TNF-α in primary infection cohorts, with attenuation or reversal of these correlations in secondary heterotypic infection [40,42]. Twelve studies paired NS1 antigenaemia with cytokines, with the strongest paired associations being NS1 with IL-10 and NS1 with IP-10; both pairings retained significance after adjustment for day of illness in three multivariable analyses [22,43]. Nine studies paired serotype with cytokine profiles, with DENV-2 showing the most pro-inflammatory profile in eastern Indian, Nicaraguan, and Vietnamese cohorts [23,44]. Seven studies paired primary versus secondary immune status with viral kinetics and cytokines, demonstrating that immune-status-modified pathogen-host interactions reshape early severity risk, particularly in adult and elderly patients [5,45]. Four studies linked endothelial markers, including angiopoietin-2 and syndecan-1, with viraemia and NS1 in the same patients, showing additive value of paired pathogen-endothelial information for plasma leakage [22,41]. A further three studies developed multivariable prediction panels combining pathogen-burden, inflammatory, endothelial, or routine clinical markers [22,46,47].

The collective signal from these paired studies is that viral burden alone is insufficient. Severity risk becomes more interpretable when viral burden is combined with immune activation, endothelial injury, and routine clinical markers, and is further modulated by immune status and infecting serotype. Selected Tier 4 prediction-model studies that simultaneously measured viraemia or NS1, a cytokine such as IL-10 or IP-10, an endothelial marker such as angiopoietin-2, and a routine chemistry such as platelet count or AST achieved internal-validation discrimination as high as an AUC of 0·96, exceeding that reported for any single domain [22,46]. This paired pathogen-host structure is the principal scientific contribution of the included evidence and the central organising message of this review.

Across paired studies, the most informative pattern was not the isolated elevation of a single cytokine or viral marker, but the alignment of viral burden, NS1 antigenaemia, cytokine amplification, endothelial activation, and routine organ-stress markers. This supports a model in which severe dengue emerges from temporally ordered pathogen-host coupling rather than from independent biomarker abnormalities.

### Quantitative synthesis of cytokine, endothelial, and pathogen-burden signatures

Quantitative synthesis was feasible for 15 biomarker domains (table 4). The main forest plot (figure 4) presents the 10 clinically prioritised markers, selected before plotting on the basis of biological plausibility, number of contributing studies, and feasibility for routine use in endemic settings. Complete pooled estimates for all 15 biomarker domains are reported in table 4. The largest pooled SMDs were observed for CXCL10/IP-10, IL-10, angiopoietin-2, AST, and ferritin; the smallest poolable effects were for quantitative viraemia and NS1 antigenaemia. Between-study heterogeneity was substantial across all domains, reflecting differences in severity definition, sampling timepoint, assay platform, immune status, and patient populations. Because pooled effects were expressed as standardised mean differences, they should be interpreted as evidence of biological direction and relative signal strength, not as clinically actionable thresholds.

**Figure 4.**
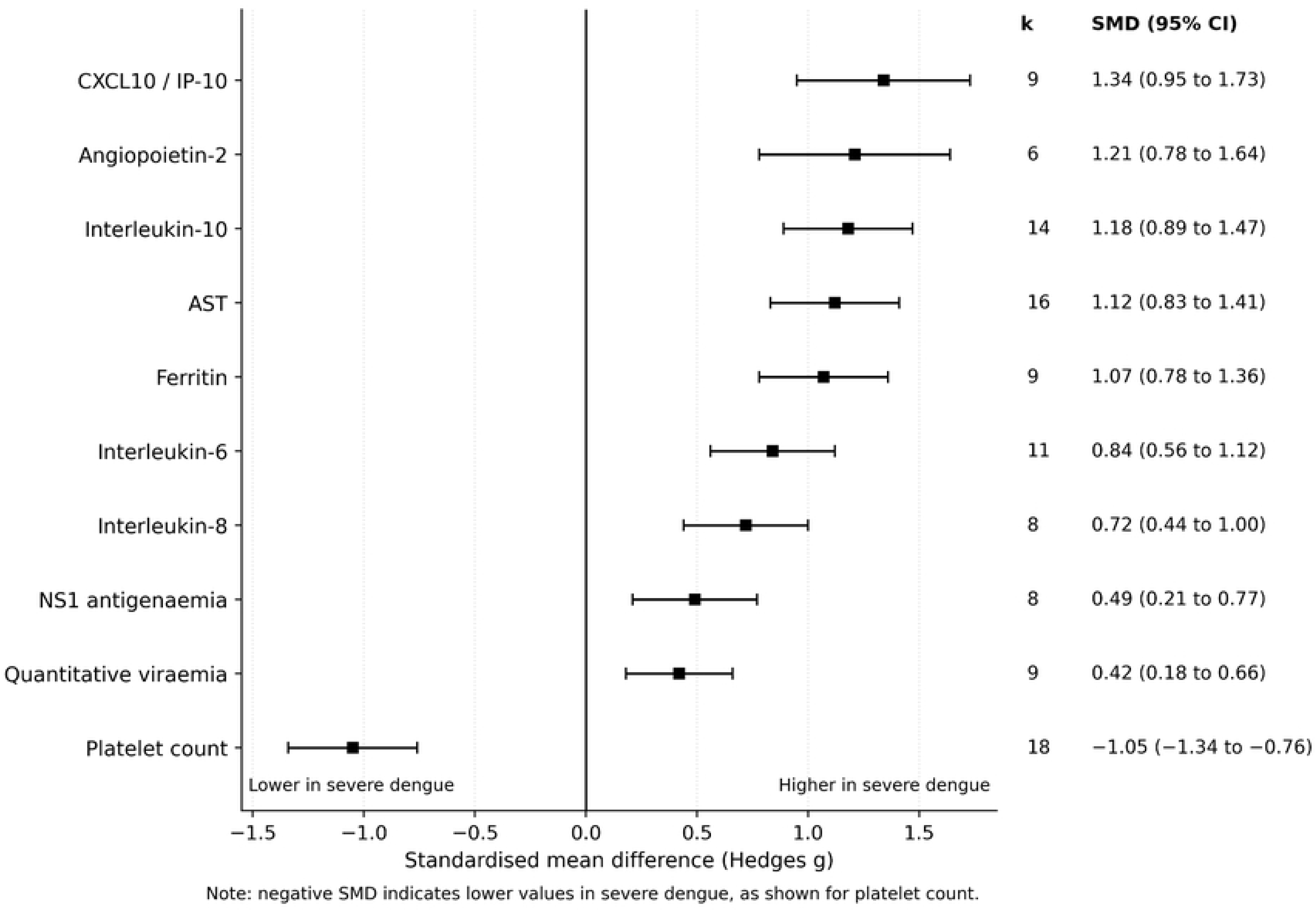
Quantitative synthesis of cytokine, endothelial, routine host-response, and pathogen-burden markers associated with severe dengue. Pooled standardised mean differences compare severe and non-severe dengue groups. Positive values indicate higher values in severe dengue, while negative values indicate lower values in severe dengue. The ten markers shown were prioritised before plotting on the basis of biological plausibility, number of contributing studies, and feasibility for routine use in endemic settings; pooled estimates for all domains are reported in table 4. Estimates should be interpreted as evidence of biological direction and relative signal strength, not as clinical thresholds.

Pre-specified subgroup analyses showed that pooled SMDs were larger in paediatric than adult cohorts for IL-10, CXCL10/IP-10, and angiopoietin-2, and larger in studies sampling during the febrile phase than during defervescence for viraemia and NS1. Studies using the WHO 2009 framework reported larger SMDs for endothelial markers than those using WHO 1997 criteria, consistent with the broader severity construct in the 2009 definition [1]. Funnel-plot asymmetry was detectable only for IL-10, suggesting possible small-study publication bias; sensitivity analysis excluding the smallest IL-10 studies attenuated the pooled estimate but did not change the direction of effect. The R script used to generate forest and funnel plot outputs is provided in Supplementary File 4.

Because pooled effects were expressed as standardised mean differences, they indicate standardised group differences rather than clinically actionable cut-offs, and should not be interpreted as bedside thresholds without further calibration in prospective cohorts.

### Translational marker panels and clinical-readiness gaps

Seven studies developed multivariable marker panels combining pathogen and host-response predictors (table 5). Reported discrimination reached areas under the curve of 0·96 in internal validation and 0·97 in discovery analyses, but the highest values came from internal or discovery settings, externally validated discrimination was lower at around 0·89, and methodological maturity was uneven. Three models were validated beyond their derivation data: a logistic-regression model combining IL-1RA, angiopoietin-2, ferritin, and additional inflammatory and endothelial markers, externally validated in a Vietnamese paediatric cohort [22]; a 20-gene transcriptomic host-response classifier with independent validation [17]; and a sex-stratified clinical bedside score based on age, comorbidities, platelet count, and transaminases, validated on a subsequent New Caledonia outbreak [47]. The remaining four models reported only internal validation by bootstrap or cross-validation. Calibration was reported in two of the models, and decision-curve analysis in none.

Sample sizes were modest, and most models contained more candidate predictors than the number of severe events would support, suggesting that some reported discrimination is likely to attenuate in external testing [36]. Models based on cytokine, endothelial, pathogen-burden, and routine clinical markers are biologically plausible, but clinical readiness remains limited by incomplete calibration reporting, sparse decision-curve analysis, and lack of validation across age groups, serotypes, immune-status strata, and endemic regions.

The prediction-model literature should therefore be viewed as a translational extension of the cytokine-endothelial evidence, not as the central result of this systematic review. Machine-learning approaches did not consistently outperform parsimonious regression models once external validation was considered. The most plausible near-term translational direction is a small, biologically interpretable panel combining one pathogen-side marker, one or two cytokine or endothelial markers, and one or two routine clinical markers. Current evidence supports development and external validation of such panels, but not routine deployment.

### Certainty of evidence

Using a structured certainty assessment informed by GRADE principles, no marker domain reached high certainty because most included studies were observational, timing-dependent, and heterogeneous in severity definition, assay platform, and statistical adjustment [51]. The pooled SMDs for IL-10, CXCL10/IP-10, AST, and platelet count were judged moderate certainty for direction of association, reflecting consistent direction across studies but substantial heterogeneity and incomplete adjustment for day of illness and immune status. Angiopoietin-2, syndecan-1, ferritin, and lactate were judged low to moderate certainty owing to smaller numbers of contributing studies. Quantitative viraemia and NS1 antigenaemia were judged moderate certainty for direction but low certainty for magnitude because their associations depend strongly on timing of sample collection. Prediction-model evidence was judged low certainty pending external validation across age groups, serotypes, immune-status strata, and endemic regions. Overall, the evidence supports mechanistic synthesis and candidate marker prioritisation rather than immediate clinical implementation.

## Discussion

This systematic review synthesises 79 studies and 47,612 participants and shows that severe dengue is best understood as a coupled pathogen-host injury process involving early viral burden, NS1-associated endothelial perturbation, cytokine and chemokine amplification, glycocalyx injury, and downstream organ stress. Across pooled analyses, the most consistent signals were not isolated markers but convergent cytokine-endothelial patterns, particularly involving IL-10, CXCL10/IP-10, IL-6, IL-8, angiopoietin-2, syndecan-1, ferritin, platelet count, and AST. Pathogen-side markers such as viraemia and NS1 antigenaemia showed more modest associations, but remain biologically important because they act upstream of immune activation and endothelial injury. The field is therefore mechanistically coherent but clinically immature.

Pathogen burden should be interpreted as an upstream trigger rather than as a stand-alone severity determinant. Viral load and NS1 antigenaemia are biologically plausible early signals because both reflect viral replication and the magnitude of endothelial exposure to viral products, but neither is uniformly associated with severity across cohorts [5,18,22,39]. Timing is central: viraemia peaks before defervescence, and studies sampling later in illness show smaller pathogen-severity associations than those sampling early in the febrile phase [5,20,21,38]. Immune status also modifies interpretation, because secondary heterotypic infection can accelerate viral clearance while amplifying immune activation through antibody-dependent enhancement and memory immune responses [10,21,28]. Serotype adds a further modifier, with DENV-2 showing the strongest inflammatory profile in several cohorts [6,23,44]. These findings support a model in which pathogen burden is clinically meaningful when interpreted together with cytokine, endothelial, and immune-status context.

NS1 deserves particular emphasis because it links viral replication to the vascular biology of severe dengue. Experimental and clinical studies show that NS1 can disrupt endothelial integrity, contribute to glycocalyx degradation, activate complement and inflammatory pathways, and promote vascular leak [18,19,29]. This makes NS1 different from a simple marker of viral presence. It is both a pathogen-burden marker and a mechanistic mediator connecting the viral compartment to endothelial dysfunction. The modest pooled severity association for NS1 in this review should therefore not be interpreted as weak biological relevance. Rather, NS1 appears to be highly timing-dependent, immune-status-dependent, and assay-dependent, with its measurable concentration shaped by viral replication, immune-complex formation, and sampling phase [21,22,39].

The host-side evidence points to a reproducible but heterogeneous cytokine-endothelial injury signature. IL-10, IL-6, IL-8, CXCL10/IP-10, ferritin, angiopoietin-2, syndecan-1, platelets, AST, ALT, and lactate all indicate immune activation, endothelial dysfunction, glycocalyx injury, or organ stress, but no single marker meets the operational criteria for a stand-alone severity test [8,11,16,22,23]. IL-10 has the largest single-marker evidence base and likely reflects immunoregulatory activation during severe disease, while CXCL10/IP-10 produced the largest pooled cytokine or chemokine effect and may capture interferon-inducible inflammatory amplification [22,23,40]. IL-6 and IL-8 showed smaller but directionally consistent effects, supporting a broader inflammatory and chemotactic profile rather than a single dominant cytokine pathway [11,23,42,43]. These findings are compatible with previous work describing severe dengue as an immune-mediated inflammatory syndrome rather than a direct consequence of viral burden alone [10,16].

Endothelial and glycocalyx markers provide the most direct biological link to the defining clinical phenotype of severe dengue: plasma leakage. Angiopoietin-2 and syndecan-1 showed strong pooled severity associations and map closely onto endothelial activation, vascular instability, and glycocalyx disruption [22,41]. This is important because severe dengue is ultimately recognised clinically through vascular leakage, haemoconcentration, shock, bleeding, and organ dysfunction [1,2,31,32]. Cytokines and chemokines may therefore be best viewed as amplifiers and regulators of injury, while endothelial and glycocalyx markers represent the vascular compartment through which severe disease becomes clinically apparent. This interpretation also explains why markers such as platelet count, AST, ferritin, and lactate retain substantial discriminatory value: they capture downstream haematological, hepatic, inflammatory, and perfusion-related consequences of the same pathogen-host injury process [7,8,22].

The central conceptual contribution of this systematic review is that severe dengue should not be framed as a single-biomarker problem. The most coherent evidence supports a parsimonious integrated panel spanning pathogen burden, cytokine or chemokine activation, endothelial or glycocalyx injury, and routine organ-stress markers. The IDAMS multi-country analysis, the largest paired-cohort study in this review, demonstrated that combinations of inflammatory and vascular markers in the febrile phase were associated with more severe outcomes and that informative marker combinations differed between children and adults [22]. Other prediction-model studies that combined viraemia or NS1, a cytokine such as IL-10 or IP-10, an endothelial marker such as angiopoietin-2, and a routine chemistry such as platelet count or AST achieved internal-validation discrimination as high as an AUC of 0·96 [22,46]. These data support integrated marker prioritisation, but not routine implementation without prospective external validation.

Translation will require deliberate attention to biological interpretability and feasibility. A useful panel must operate during the febrile phase, before warning signs and plasma leakage are clinically established; it must remain robust across age groups, serotypes, and immune-status strata; and it must be feasible in endemic settings where multiplex cytokine or endothelial assays may not be routinely available [1,8,9]. A practical pathway may involve anchoring widely available markers, such as NS1 antigenaemia, platelet count, AST, and ferritin, to one or two more specific cytokine or endothelial markers, such as IL-10, CXCL10/IP-10, angiopoietin-2, or syndecan-1. The externally validated models identified in this review provide an instructive template, but none should yet be considered ready for routine clinical use without further prospective testing across regions and care settings [17,22,47].

The prediction-model literature should be interpreted as a translational extension of the cytokine-endothelial evidence, not as the central finding of this systematic review. Reported discrimination reached AUCs of 0·96 in internal validation and 0·97 in discovery analyses, but the highest values came from internal or discovery settings with few severe events, calibration was inconsistently reported, decision-curve analysis was rare, and most models lacked external validation. These issues are important because models with strong internal discrimination may perform less well when applied to new populations, especially when severe-event counts are small, predictors are numerous, or sampling time differs between cohorts [36,49]. Machine-learning approaches did not consistently outperform parsimonious regression models once external validation was considered. The more defensible near-term direction is therefore a small, interpretable, biologically grounded marker panel rather than a complex high-dimensional prediction algorithm.

Future studies should be designed around the pathogen-cytokine-endothelial axis rather than around isolated biomarkers. Prospective cohorts should record exact day of illness at sampling, infecting serotype, primary or secondary immune status, viral load or NS1 assay platform, cytokine and endothelial assay methods, severity definitions, group sizes for severe and non-severe categories, adjusted effect estimates with confidence intervals, and internal and external validation of any proposed marker panel. Reporting standards for prognostic-factor studies and prediction models should be applied consistently [48,49]. Raw data sharing should accompany publication where possible, because lack of extractable group-level summary statistics remains a major barrier to quantitative synthesis. The next research priority is prospective external validation of affordable early-phase cytokine-endothelial-pathogen panels across age groups, serotypes, immune-status strata, and endemic regions.

The strengths of this systematic review include prospective PROSPERO registration, conduct and reporting in accordance with PRISMA 2020, a comprehensive multi-database search without language restriction, integrated assessment of pathogen-related and host-response evidence in a single synthesis, design-appropriate risk-of-bias evaluation using QUIPS, PROBAST, JBI, and selective NOS cross-walks, and quantitative synthesis where data permitted [33,35,36,37,50]. The review is, to our knowledge, the first to align severe dengue biomarker evidence directly with a pathogen-cytokine-endothelial framework rather than treating viral kinetics, cytokines, endothelial markers, and clinical markers as separate literatures.

Several limitations should be acknowledged. Between-study heterogeneity in severity definitions, sampling time, assay platforms, immune-status classification, and statistical adjustment was substantial across nearly every biomarker domain, limiting the precision of pooled estimates. Severe-disease group sizes were modest in most cohorts, increasing the influence of small-study bias and constraining subgroup analysis. Hospital-based recruitment in most studies introduces selection bias towards clinically apparent disease and limits generalisability to community-managed cases. African and Pacific data were sparse, limiting inference in regions where dengue surveillance and biomarker infrastructure remain underdeveloped. Several relevant studies could not be included in pooled analysis because group-level summary statistics were unavailable. Inconsistent reporting of adjustment variables and incomplete external validation of marker panels limit conclusions about clinical translation. These limitations mean that the evidence supports mechanistic synthesis and candidate marker prioritisation, but not immediate clinical implementation.

In conclusion, severe dengue is most coherently understood as an integrated pathogen-host injury syndrome. Viral burden and NS1 antigenaemia provide early upstream signals, while cytokine and chemokine amplification, endothelial activation, glycocalyx injury, and routine markers of organ stress capture the host processes that drive severe clinical phenotypes. The current evidence supports prospective validation of parsimonious early-phase cytokine-endothelial-pathogen panels, but does not yet support routine clinical deployment. External validation across age groups, serotypes, immune-status strata, and endemic regions is required before clinical implementation can be recommended.

## Data Availability

All data underlying the findings of this study are provided within the main manuscript and the accompanying supplementary files.

## Contributors

PMA conceived and designed the review, registered the PROSPERO protocol, and designed the search strategies. PMA and PA independently screened titles, abstracts, and full texts, jointly performed data extraction, and verified inclusion decisions. PA independently verified risk-of-bias judgements for randomly selected subsets of the included studies. AK and VK contributed methodological input on systematic review conduct and prognostic-factor synthesis. LA contributed to manuscript drafting and revision. PMA performed the statistical analyses, drafted the manuscript, and integrated revisions. All authors reviewed and approved the final version. PMA had full access to all data in the study and accepts final responsibility for the decision to submit for publication.

## Data sharing

The full search strategies and PRISMA 2020 checklist are provided in Supplementary File 1; the screening and study-decision log is provided in Supplementary File 2; the risk-of-bias sheets (QUIPS, PROBAST, JBI, and the selective NOS cross-walk where applicable) are provided in Supplementary File 3; and R scripts used for meta-analysis are provided in Supplementary File 4. De-identified individual study data extracted from published reports will be available with the manuscript at the journal website upon publication.

## Acknowledgements

We thank the corresponding authors of included studies who provided clarifications and unpublished group-level summary statistics in response to email queries.

You can use this in the manuscript, usually after **Declaration of interests** or before **Acknowledgements.**

## Generative artificial intelligence statement

Generative artificial intelligence tools were used only for language editing, grammar correction, formatting assistance, and improving clarity of expression during manuscript preparation. The tools were not used to generate original scientific data, perform study selection, extract data, assess risk of bias, conduct statistical analyses, or make decisions regarding interpretation of results. All AI-assisted text was critically reviewed, edited, and approved by the authors, who take full responsibility for the accuracy, integrity, and final content of the manuscript.

